# Building a Roadmap for Nutrition Education in Medical Training: Lessons from a Student-Led Pilot

**DOI:** 10.1101/2025.11.03.25339436

**Authors:** Akash Patel, Ritika Modi, William McGonigle, Gauri Agarwal

## Abstract

**Background:** Poor diet is the leading cause of global disability and premature death, contributing to 11 million deaths annually. Despite this, nutrition remains underemphasized in medical education, with over 70% of United States (U.S.) medical schools failing to meet the National Research Council’s 1985 recommendation of 25 hours of nutrition training. In 2022, the U.S. House of Representatives passed a bipartisan resolution calling for meaningful nutrition education for health professionals. Amid this gap, unreliable media sources frequently shape patient nutrition knowledge rather than professional sources such as registered dietitians or licensed physicians. Strengthening interprofessional collaboration with nutrition professionals could enhance dietary counseling and patient care.

**Methods:** In response, one student implemented a pilot three-part clinical nutrition lecture series at the University of Miami Miller School of Medicine (UMMSM). Resulting student enthusiasm catalyzed the formation of a 50-member taskforce formed to expand and integrate nutrition longitudinally. A pre- and post-lecture survey around the first of three planned sessions (Npre = 98, Npost = 77) assessed knowledge, perceptions of nutrition training, and comfort with dietary counseling.

**Results:** Knowledge of evidence-based nutrition improved significantly (p < .001). Post-lecture, students reported greater confidence applying nutrition in clinical practice and increased interest in lifestyle medicine training (p < .001).

**Conclusions:** The first session enhanced students’ practical skills and understanding of nutrition’s role in health. This trajectory illustrates how even a small pilot can stimulate sustainable reform. We discuss key elements of an effective, multifaceted nutrition curriculum and propose a roadmap adaptable to other institutions.

## Introduction

Suboptimal diet is a leading contributor to global morbidity and mortality, associated with an estimated 11 million deaths and 255 million disability-adjusted life years (DALYs) worldwide in 2017 alone [1]. Poor dietary patterns are a major risk factor for noncommunicable diseases such as cardiovascular disease, type 2 diabetes, and certain cancers. In the U.S., dietary risks represent the largest individual risk factor for mortality, surpassing tobacco use, obesity, and other commonly cited health risks [2, 3]. There exists a well-established association between unhealthy dietary patterns and reductions in both functional ability and life expectancy across populations [4].

Despite the clear impact of nutrition on health outcomes, medical education in the U.S. continues to underemphasize nutrition and dietary counseling. As early as 1985, the National Research Council recommended that medical schools provide at least 25 hours of nutrition education, yet over 70% of U.S. programs have failed to meet this benchmark. [5, 6] At the same time, growing public interest in diet and preventive health has led to increased patient demand for accurate, evidence-based nutritional guidance [7].

Undergraduate medical education in the U.S. is accredited by the Liaison Committee on Medical Education (LCME), which does not require minimum hours of nutrition training or mandate applied instruction in counseling or chronic disease management [8]. Consequently, the amount and focus of nutrition teaching varies widely across schools. Much of the nutrition content traditionally encountered by medical students is biochemistry-focused (micronutrient metabolism and deficiency syndromes) but does not prioritize clinically oriented skills in counseling or diet-related chronic disease management [9]. Surveys consistently show insufficient preparation for evidence-based nutrition counseling and limited opportunities to practice applied skills [10, 11].

Some medical schools have introduced nutrition electives, integrative medicine modules and teaching kitchen experiences [12]. However, these opportunities are often optional and inconsistently available, leaving many students without exposure to foundational nutrition training. Even when available, elective offerings vary widely in content, quality, and duration across institutions [9]. In a 2025 national survey, the Association of American Medical Colleges found that although 100% of schools included some nutrition in required curricula, fewer than half (45%) extended it across multiple courses, and only 17% achieved longitudinal integration across all years [13].

Expert groups and recent reviews have repeatedly noted this gap and called for greater standardization [10]. The recent JAMA consensus statement proposes 36 nutrition competencies and explicitly maps them to the Accreditation Council for Graduate Medical Education (ACGME) six core competency domains to facilitate integration across undergraduate and graduate training [14]. Recognizing this gap, the U.S. House of Representatives passed a bipartisan resolution in 2022 (H.Res.1118), calling for more comprehensive nutrition education for health professionals [14].

The American Heart Association (AHA) has echoed these concerns, noting that physicians without adequate nutrition training often lack the confidence to assess or counsel patients on dietary issues [11]. As a result, patients increasingly turn to unregulated media and social platforms for nutrition advice, sources that may propagate misinformation or even harmful guidance. The AHA advocates for the integration of longitudinal, systems-based nutrition education throughout medical curricula, with an emphasis on clinical relevance and interprofessional collaboration [15]. These recommendations also include structured learning alongside registered dietitians to promote both knowledge acquisition and appropriate referral practices.

In a wider context, it is important to acknowledge that improving physician training is only one part of the broader solution. Meaningful progress will also depend on changes to the food environment, including policies that expand access to affordable healthy foods, industry reforms, and stronger integration of registered dietitians into patient care. Expanding insurance coverage for medical nutrition therapy and establishing referral pathways are essential system-level needs. Within this wider lens, medical schools nonetheless have a clear and actionable opportunity to improve training.

In response to the recognized deficiency of formal nutrition education, one student worked with faculty to design and deliver a three-part lecture series spanning the cardiology, gastroenterology, and endocrinology blocks. Its success spurred the formation of a student-led taskforce that is now collaborating with faculty to embed nutrition longitudinally at UMMSM. This study aimed to evaluate a student-led pilot nutrition education initiative and to propose a framework for longitudinal, competency-based integration of nutrition across medical training.

## Methods

We conducted a pre-post intervention study to evaluate the impact of a clinical nutrition lecture series implemented at a single center during the 2023 academic year.

The lecture series was designed and delivered by one student in collaboration with faculty. It was structured as a systems-based, clinically oriented series integrated into existing organ system courses, including Cardiology, Gastroenterology, and Endocrinology.

Each 45-minute interactive lecture covered evidence-based nutrition principles relevant to the specific organ system, including macronutrient and micronutrient fundamentals, diet-related chronic disease management, and practical strategies for patient dietary counseling. All lecture content was reviewed and approved by respective course directors for accuracy, clinical relevance, and integration with existing curricula.

The pilot cardiovascular nutrition lecture was delivered in-person to first-year medical students in October 2023. The session employed active learning techniques including case-based discussions, interactive polling, and small group exercises focused on dietary assessment and counseling scenarios.

Evaluation included a knowledge assessment and self-reported competency, attitudes, and session evaluation utilizing a Likert scale. Pre-lecture surveys were administered electronically immediately before the first nutrition session and post-lecture surveys were distributed via QR code after the first lecture. All responses were anonymous and linked using a unique, self-generated identifier code.

All first-year medical students (n=192) who attended the lecture were eligible to participate. Students were informed that participation in the survey evaluation was voluntary and would not affect their course grades.

Due to non-normal distribution of Likert scale responses (confirmed by Shapiro-Wilk tests, p < 0.05), non-parametric analyses were employed. Mann-Whitney U tests were used to compare pre- and post-survey responses between groups, as matched pairs could not be reliably established due to the anonymous survey design and variable response rates. Statistical significance was set at p < 0.05. Analyses were performed with IBM SPSS Statistics, version 29.0. No a priori power calculation was performed given the pilot evaluation design. No adjustments for multiple comparisons were applied; results are interpreted as exploratory. Analyses were conducted on available cases; no imputation was used for missing responses. All pre- and post-session survey data are available in the Supporting Information (*S1 Data*). This study was approved by the University of Miami Institutional Review Board (Protocol #20231027) and determined to be exempt under 45 CFR 46.104. Recruitment and data collection occurred on October 23, 2023, coinciding with delivery of the cardiovascular nutrition lecture. All participants provided written informed consent electronically prior to participation through a digital form embedded in the pre-lecture survey. Participation was voluntary, no identifying information was collected, and responses were analyzed in aggregate.

## Results

Of the 192 eligible first-year medical students, 98 completed the pre-lecture survey (response rate: 51%) and 77 completed the post-lecture survey (response rate: 40%). The overall matched response rate, accounting for students who completed both surveys, was 39% (n=75). Knowledge assessment scores improved significantly (Med_pre_ = 6, Med_post_ = 8; U = 862.00, Z = −8.622, p < .001), underscoring the effectiveness of a focused, evidence-based nutrition intervention.

Self-reported knowledge of evidence-based nutrition increased significantly (Med_pre_ = 2, Med_post_ = 3; U = 1883.00, Z = −5.694, p < .001). Confidence in applying nutrition principles to patient care also improved significantly (Med_pre_ = 2, Med_post_ = 3; U = 1702.50, Z = −5.847, p < .001).

No statistically significant change was observed in perceived value of nutrition in disease management (Med_pre_ = 4, Med_post_ = 4; *U* = 3377.50, *Z* = –1.780, *p* = .075) or in perceived confusion/contradiction in the nutrition space (Med_pre_ = 4, Med_post_ = 4; *U* = 3636.50, *Z* = –0.170, *p* = .865). However, interest in the inclusion of more lifestyle medicine in medical education increased significantly (Med_pre_ = 4, Med_post_ = 4; *U* = 2779.50, *Z* = –3.583, *p* < .001).

Pre-survey, most respondents reported receiving at least some nutrition information from social media, with 31.63% indicating a moderate amount, 30.61% a little, 11.22% a great deal, and 20.41% none at all.

Post-survey only items indicated that participants rated the session as moderately to highly influential on their own lifestyle choices (Mean 3.26/4), likely to influence their future patient counseling (Mean = 3.52/4), and valuable overall (Mean 3.8/4).

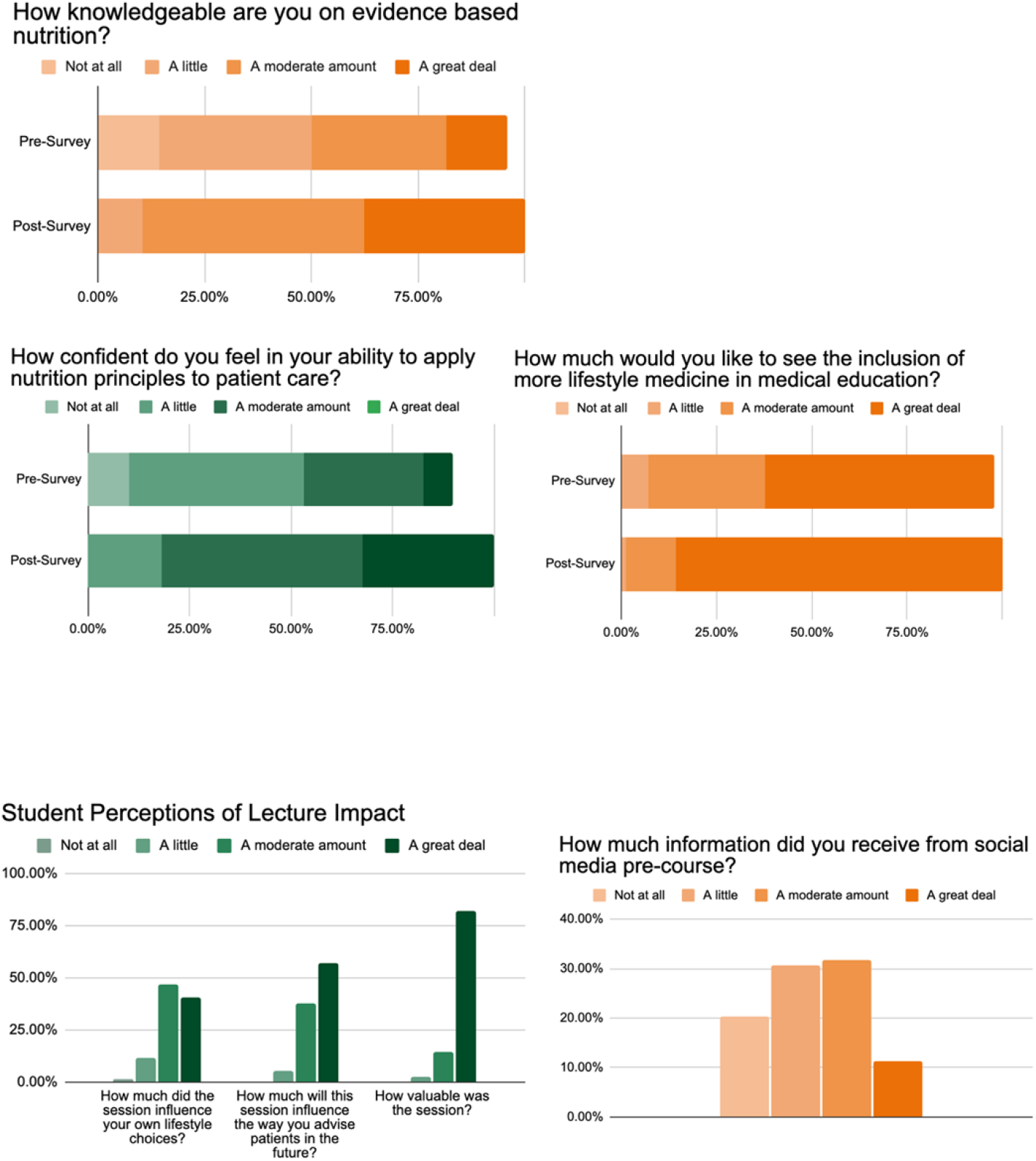

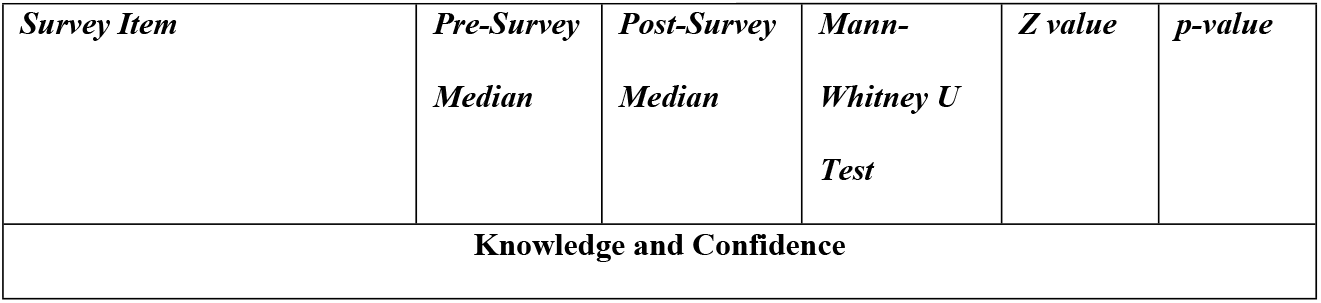
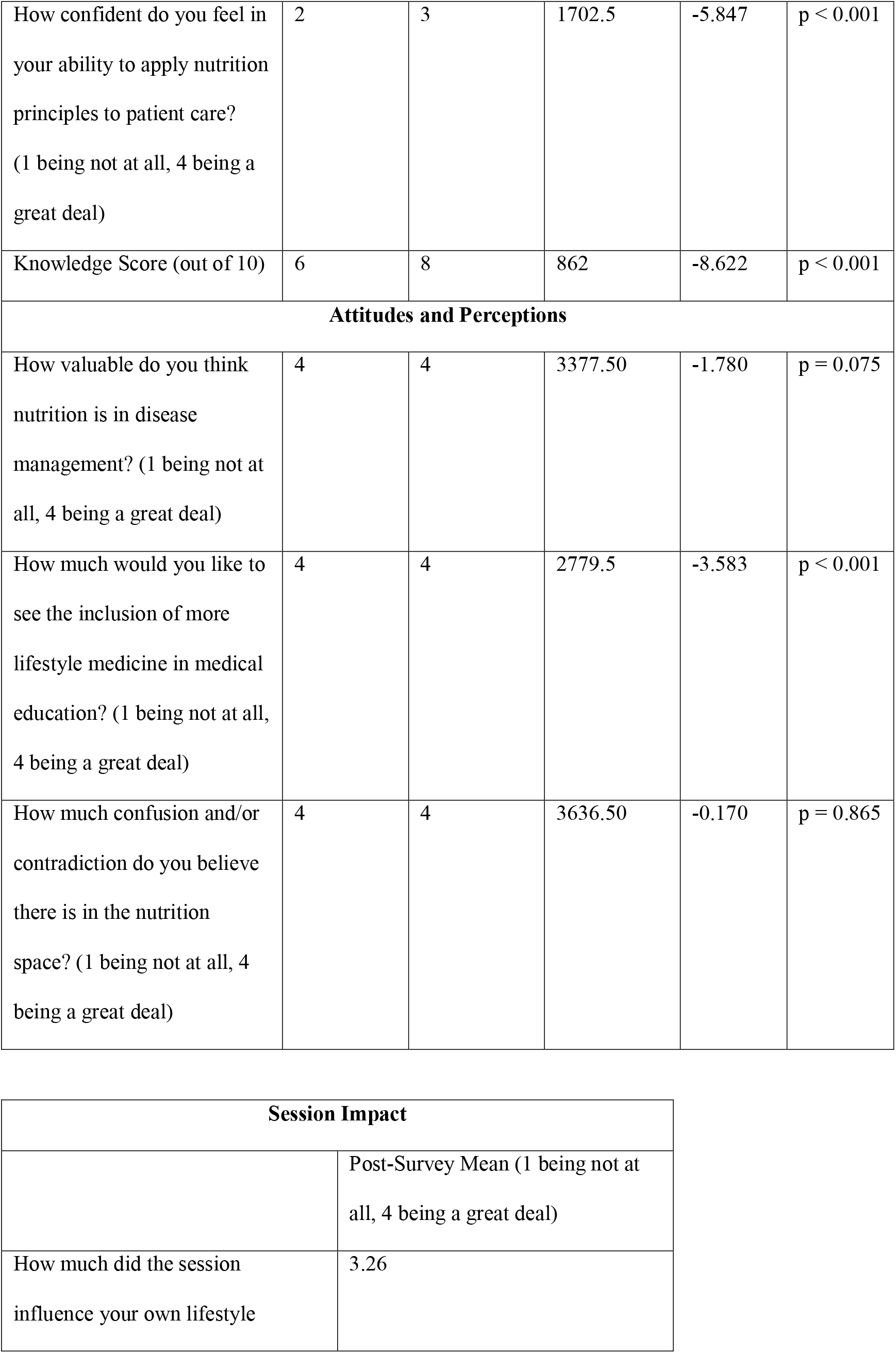
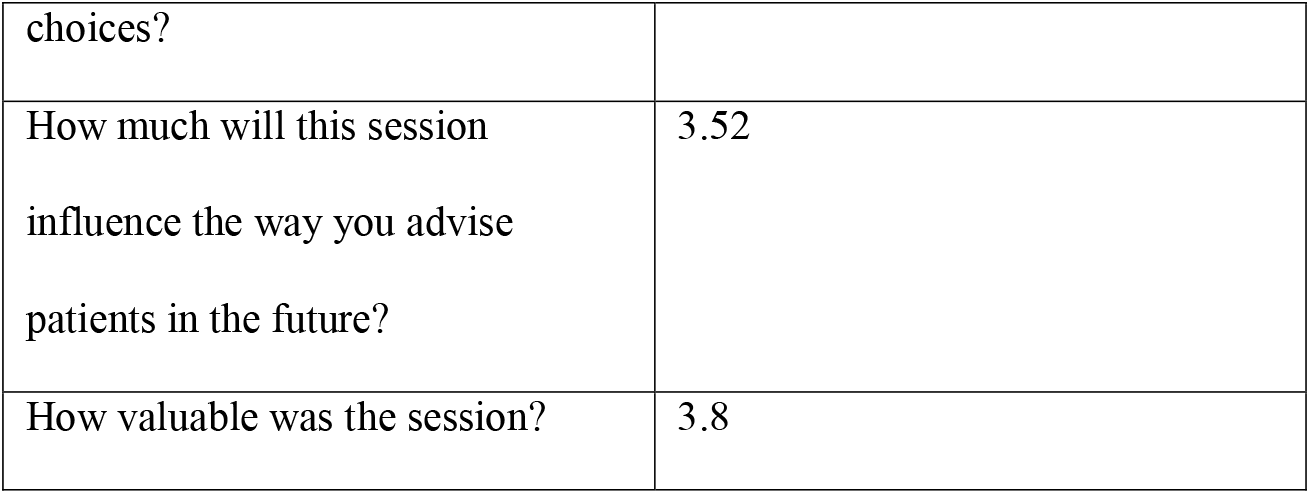

## Discussion

### Impact of the Pilot Lecture: Catalyzing Institutional Change

Findings from the first lecture in this pilot series demonstrate that even a single, targeted session can meaningfully improve medical students’ nutrition knowledge and confidence. More importantly, the pilot sparked momentum for broader change. Following overwhelmingly positive feedback, faculty invited the lecture to be repeated and expanded into other modules, and the success of the lecture series, combined with strong enthusiasm from peers, led to the formation of a 50-member student taskforce committed to advancing nutrition education. This taskforce now collaborates with faculty to design a longitudinal, systems-based curriculum at the UMMSM and is developing resources adaptable for medical students and health professionals nationally. These findings demonstrate that the pilot achieved its aim of improving student knowledge and confidence while creating momentum for longitudinal curriculum reform.

These developments highlight how a small, student-led pilot can catalyze institutional reform when supported by faculty engagement and learner demand. At the same time, isolated lectures remain insufficient to close the gap in medical nutrition education and translate to improved public health. A lasting solution requires a curriculum that is mandatory, sustainable, and longitudinally integrated.

### Implementation Options for Nutrition Education

The recent JAMA consensus statement identifies 36 nutrition competencies for medical trainees and maps them to the ACGME’s six core competency domains (patient care and procedural skills; medical knowledge; practice-based learning and improvement; interpersonal and communication skills; professionalism; and systems-based practice). Building on that framework, we propose a multifaceted, longitudinal model that operationalizes these competencies across the curriculum [14]. While several approaches occur concurrently rather than sequentially, layering them from pre-clinical through graduate medical education ensures competencies are introduced, reinforced, and applied in multiple clinical contexts. At UMMSM, this has taken shape through the following complementary strategies:

#### Pre-Learning Resources

Foundational content can be delivered before class through concise, standardized resources. For example, self-paced online modules have proven effective in graduate medical education and can be completed independently and assessed internally [16]. In parallel, the UMMSM taskforce is developing short, interactive digital articles widely used in medical education, to introduce targeted nutrition concepts before in-class discussions [17]. *Primarily advances Medical Knowledge and Practice-Based Learning and Improvement* [14].

#### Lectures and Live Teaching

Didactic sessions remain valuable for establishing frameworks and highlighting high-yield topics. Their effectiveness is greatest when paired with active elements such as polling or small group discussion. The UMMSM pilot lecture series, which began in cardiology and expanded into gastroenterology and endocrinology, illustrates how lectures can serve as effective entry points for broader curricular reform. *Primarily advances Medical Knowledge; when interactive, also supports Practice-Based Learning and Improvement* [14].

#### Case-Based Learning (CBL)

CBL is already a core pedagogy in most U.S. medical schools, where students collaboratively analyze patient cases to build clinical reasoning. Embedding nutrition content into these cases normalizes dietary counseling as part of standard patient management. At UMMSM, nutrition questions are being incorporated into CBL scenarios for conditions such as diabetes and hyperlipidemia. *Primarily advances Patient Care and Procedural Skills and Practice-Based Learning and Improvement* [14].

#### Standardized Patients

Working with standardized patients (actors trained to simulate clinical encounters) provides a safe environment for students to practice motivational interviewing and personalize nutrition recommendations. These sessions also allow for structured feedback on counseling skills, though they require additional faculty time and institutional resources. *Primarily advances Interpersonal and Communication Skills and Professionalism; secondarily Patient Care* [14].

#### Teaching Kitchens

Teaching kitchens provide experiential, hands-on opportunities for students to learn nutrition through cooking and meal preparation. This model combines didactic instruction with practical skills, reinforcing concepts of healthy eating, food literacy, and patient counseling. At UMMSM, a pilot culinary medicine elective has been developed to introduce students to evidence-based nutrition principles in an applied setting, while also highlighting strategies for patient engagement. Although resource-intensive, teaching kitchens can foster collaboration with dietitians and chefs and are increasingly recognized as a scalable modality for nutrition education. *Primarily advances Interpersonal and Communication Skills and Professionalism; secondarily Patient Care* [12, 14].

#### Graduate Medical Education

Nutrition training should extend beyond medical school. Specialty-specific content in residency and fellowship ensures that nutrition is reinforced when it is most clinically relevant and allows tailoring to specialty populations (e.g., cardiovascular disease, cancer survivorship, or chronic kidney disease). *Primarily advances Systems-Based Practice* [11, 14].

These components are most impactful when layered together, and each advances different domains within the ACGME core competency framework. In Figure 1, we annotate every implementation option with the primary ACGME domain(s) it addresses to show how the model operationalizes the JAMA consensus nutrition competencies across the curriculum [14].

**Fig. 1.**
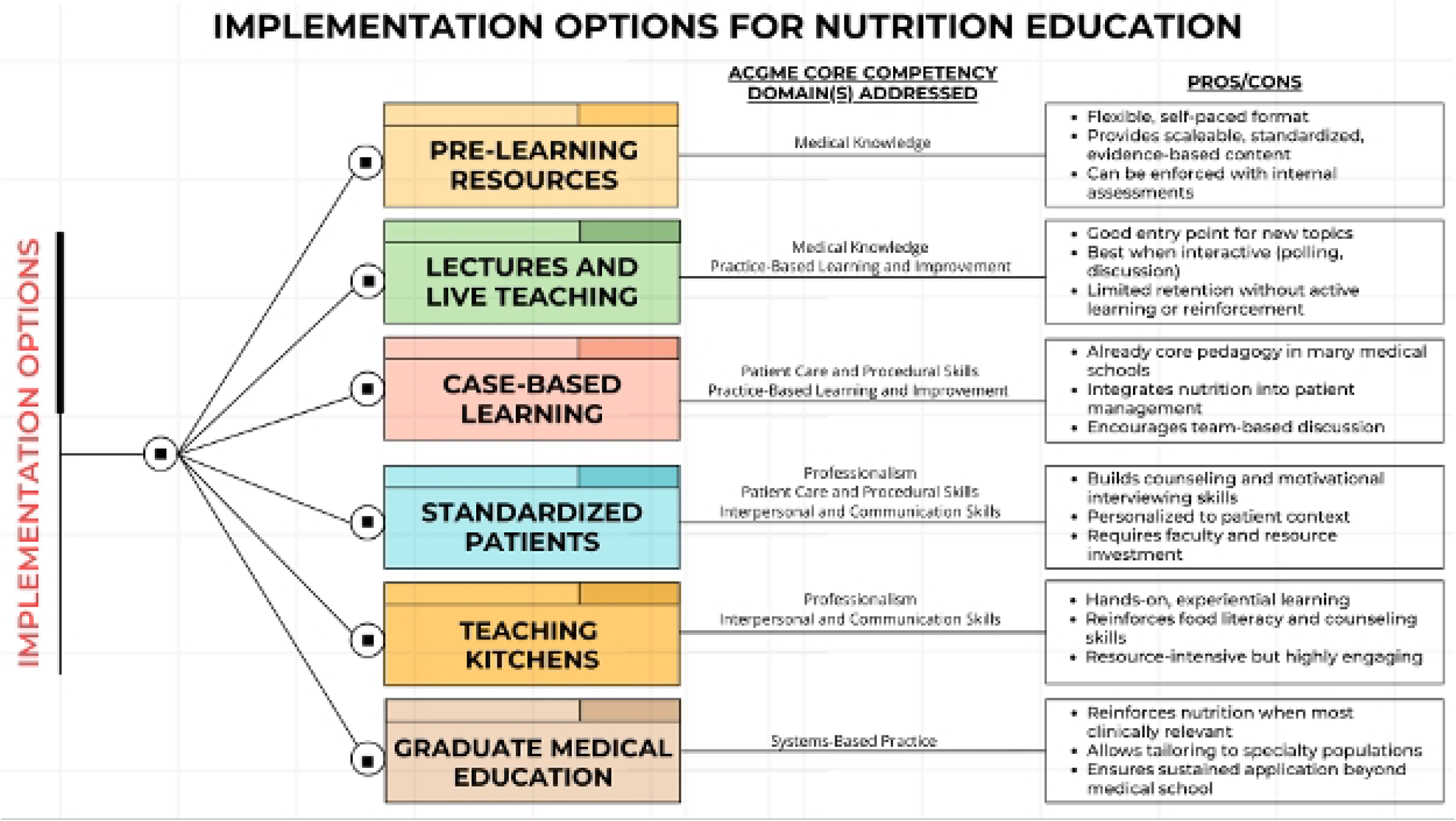
Implementation options for nutrition education and the ACGME competency domains they address. Pathways include pre-learning resources, lectures and live teaching, case-based learning, standardized patients, teaching kitchens, and graduate medical education. Each approach lists the primary ACGME domain(s) addressed (Medical Knowledge; Patient Care and Procedural Skills; Practice-Based Learning and Improvement; Interpersonal and Communication Skills; Professionalism; Systems-Based Practice) alongside illustrative pros/cons. Approaches may occur concurrently rather than sequentially; layering them across training reinforces and applies nutrition competencies in diverse contexts

### Towards Standardization and Broader Adoption

Wider adoption across institutions will ultimately depend on standardization. The recently proposed core competencies in nutrition education offer a roadmap for curriculum design, evaluation, and faculty development. Since these competencies are categorized within the ACGME’s six core domains, linking implementation strategies and assessments to those domains can streamline integration with existing milestones, entrustable professional activities (EPAs), and program evaluation [14]. National medical education organizations and accrediting bodies can help promote consistency and support quality control across institutions. Aligning implementation strategies with recent policy trends may facilitate broader uptake and sustained momentum. The AAMC report highlighted that only 17% of medical schools achieve longitudinal integration, underscoring the urgency of operationalizing nutrition competencies through structured, multi-modal strategies [13].

### Limitations

This study evaluated a single lecture at a single institution with voluntary participation, modest response rates, and anonymous surveys that prevented reliable matching of pre- and post-responses. Outcomes were short-term and largely self-reported. Results may be influenced by response bias and may not generalize to other settings. Future work should include multi-site pilots, longitudinal retention and patient-level outcomes.

### Role of Physicians and Dietitians

While registered dietitians are the recognized experts in nutrition care and should be prioritized as central members of the healthcare team, there remain systemic barriers to fully leveraging their expertise. Coverage for medical nutrition therapy is limited, referral pathways are inconsistent, and dietitians are not available in all care settings [11]. In this context, equipping physicians with foundational nutrition training is not intended to supplant the role of dietitians, but rather to complement and amplify it. Physicians remain the first point of contact for most patients, and as central coordinators of care, they have a unique opportunity to influence patient perceptions and behaviors. Even brief, evidence-based nutrition guidance from physicians can help normalize dietary counseling as part of standard medical care and can facilitate timely referrals to dietitians. Moreover, physician training in nutrition may strengthen advocacy for expanded access to dietitian services. Importantly, any nutrition education added to the medical curriculum must be carefully considered within the already crowded landscape of medical training, continued throughout residency and fellowship where it has the greatest clinical relevance, and focused on the practical skills needed to work effectively alongside dietitians and other professionals.

## Conclusion

Strengthening physician training in nutrition is a necessary step toward addressing the growing burden of diet-related disease. While our findings are limited to the first lecture in a three-part series, the impact extended beyond measurable knowledge gains: faculty implemented additional nutrition objectives and student enthusiasm led to the creation of a 50-member taskforce dedicated to further curriculum reform. This illustrates how even a small pilot can evolve into a sustainable, multifaceted student-driven model for institutional change. With institutional support and alignment with national competencies, medical education can begin to close the gap between what patients need and what future physicians are prepared to deliver. Such training should be viewed as complementary to, not a replacement for, the expertise of registered dietitians. Equipping physicians with baseline skills can also help strengthen collaboration and advocacy for expanded access to dietitian services.

## Statements and Declarations

### Human Ethics and Consent to Participate

This study was approved by the University of Miami Institutional Review Board (Protocol #20231027) and conducted in accordance with the principles of the Declaration of Helsinki. All participants provided written informed consent prior to participation through a digital form embedded in the pre-lecture survey. Participation was voluntary, and no identifying information was collected. The study followed the STROBE guidelines for reporting observational studies.

### Data Availability

All de-identified data underlying the findings are provided in the Supporting Information file *S1 Data*.

### Author Contributions

A.P. conceived the curriculum, developed the study design, and led the manuscript concept. G.A. provided oversight, approved curriculum development, and supervised the project and manuscript preparation. R.M. conducted the data analysis. W.M. contributed to drafting and revising the manuscript. All authors reviewed and approved the final version.

## Acknowledgments

We are grateful to Dr. Michael Dyal (Cardiology), Dr. Siobhan Proksell (Gastroenterology), and Dr. Diana Soliman (Endocrinology) for their support as course directors in allowing delivery of the nutrition lectures within their modules. We also thank the administration of the University of Miami Miller School of Medicine for their receptiveness and support in advancing these curricular changes. All individuals acknowledged have provided consent to be named.

## Notes

### Competing Interest Statement

The authors have declared no competing interest.

### Funding Statement

The author(s) received no specific funding for this work.

### Author Declarations

University of Miami Institutional Review Board

### Summary of Updates

Uploaded clean manuscript to remove visible markup bar. No content changes.

